# Relationship dynamics and behavioral adaptations in the control of the 2022 mpox epidemic

**DOI:** 10.1101/2025.05.02.25326849

**Authors:** Ulrik Hvid, Lone Simonsen, Morten Frisch, Kim Sneppen

**Author notes:** U.H., K.S. and L.S. designed research; U.H. and K.S. performed research; U.H. and K.S. analyzed data; and U.H., L.S. and K.S. wrote the paper. M.F. provided the Project SEXUS data. All authors critically revised the manuscript. The authors declare no competing interest.

## Abstract

We analyzed the patterns of transmission in the 2022 mpox epidemic as it unfolded in the European population of MSM (men who have sex with men). We developed an agent-based model that simulates sexual pair formation, incorporating both brief and longer-term sexual relationships. The model implements survey data on the sexual behavior of MSM and accounts for the highly heterogeneous nature of the sexual contact network within this community. When simulating the mpox epidemic, the model reproduces the reported numbers of sexual partners of mpox-infected patients. We find that natural herd immunity had little impact on ending the European outbreak. Instead, we suggest that the marked decrease in serial interval observed across the epidemic reflects a dramatic increase in self-isolating behavior amongst infected and that this is sufficient to explain the early control of the epidemic. Our work highlights the critical interplay between relationship dynamics and adaptive behaviors in shaping mpox epidemic patterns and achieving control in 2022. Now that the virus is endemic, the European MSM population remains protected by a combined effect of increased awareness and immunity, both natural and vaccine-induced.

**Significance Statement:** The waning of smallpox immunity since its elimination around 1980 leaves an immunological opening to emerging poxviruses. We have witnessed two major mpox outbreaks – clade IIb in 2022 and the ongoing clade Ib in Central Africa – both concentrated in a subpopulation with high-risk sexual behavior. Clade IIb is now endemic and recently surged among Australian MSM. Despite parallels with HIV, particularly its disproportionate burden on MSM, the short infection cycle and subsequent lifelong immunity of mpox imply distinct transmission dynamics. These novel outbreaks demand new modeling approaches and a deeper grasp of herd immunity in highly heterogeneous sexual networks.

---

**M**pox (formerly monkeypox) clades I and II have caused zoonotic outbreaks in central and western Africa for many decades. In the spring of 2022, however, a newly emerged variant of clade II, named IIb, spread from Nigeria to Europe and subsequently caused 100,000 mpox cases globally (1). The virus may have been circulating unnoticed in Europe for months (2), before spreading explosively in the aftermath of a few superspreader events, most significantly the Maspalomas Gay Pride in Gran Canaria (3). The epidemic was overwhelmingly concentrated in men who have sex with men (MSM) (4). Furthermore, the observed HIV comorbidity rate of 38% in Europe (4) strongly suggests that the epidemic was mainly driven by, and concentrated in, a “core group” with a high-risk sexual lifestyle.

In Europe, the epidemic peaked in early July, and the Americas followed soon after (5). At that time, less than 15,000 cases were registered in Europe, corresponding to roughly 0.2% of the MSM population there. Several studies have proposed different explanations as to what caused this decline. In Europe, the IMVANEX vaccine was approved on July 22 (4), and in most countries widespread pre-exposure vaccination began as late as mid-August, well after the reproduction number had gone below 1 (Fig. 1 B). Thus, vaccination efforts can be ruled out as an explanation for the waning epidemic. Some have pointed to immunity in the core group as the main driver of control (6)(7)(8). Others have suggested that it was a decrease in sexual activity among the core group that caused the decline (9)(10)(11), together with a decreased effective infectious period due to increased awareness of symptoms (10). The hypothesis of decreased sexual activity is challenged by two empirical studies which found no decrease in other sexually transmitted infections (STIs) among HIV-PrEP users - a proxy for the core group - during the mpox epidemic (12)(13). However, the significant decrease in the mean serial interval for mpox infections observed during the epidemic in several countries may be attributed to infected individuals abstaining from contact during their infectious period (10). We will use the term “responsive behavior change” to describe a change in which people become more likely to decrease risk behavior *after* acquiring symptoms, minimizing risk to others, as opposed to a “preventive behavior change”, where people decrease risk behavior irrespective of symptoms, to protect themselves from infection.

**Fig. 1.**
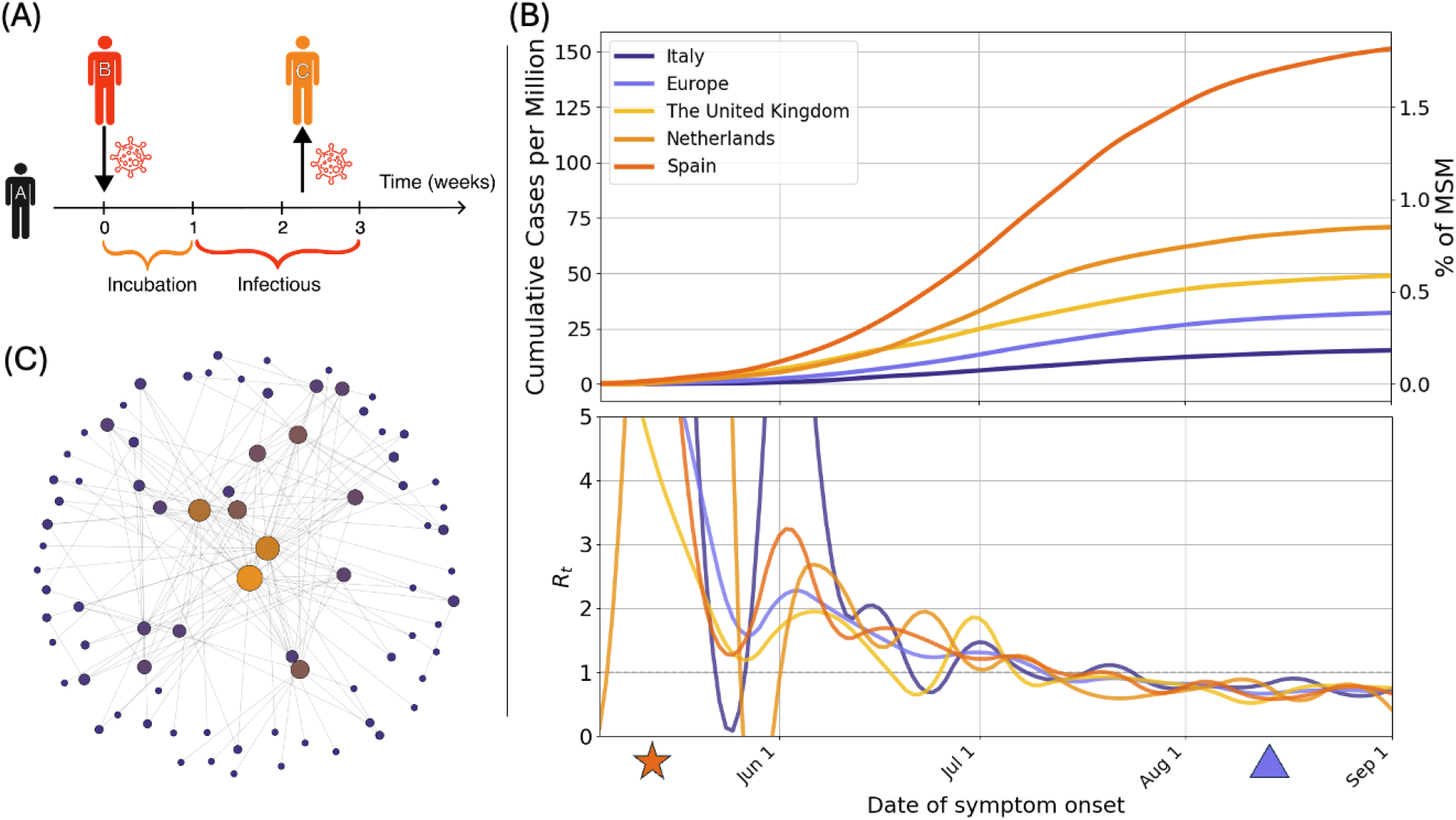
European Clade IIb mpox outbreaks and transmission dynamics in the MSM network. **(A)** Cartoon of incubation and infectious period in a typical mpox infection cycle. At least two unique sex partners within the ≈ 3 week period from infection to viral clearance and immunity are needed for a sustained epidemic. **(B)** Top, cumulative mpox cases per million persons in European countries during summer 2022 shows substantial country-to-country variation. Bottom observed reproduction numbers *R*_*e*_ in these countries, estimated based on serial interval and the ratio between cases reported in consecutive weeks; One observes a decline in *R*_*e*_ over time, with variation associated with large randomness in the early weeks. The timing of the Maspalomas Gay Pride is indicated with 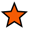 and the approximate start of the PEP vaccination rollout marked with 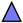. **(C)** An illustration of the highly heterogeneous MSM network based on sexual activity is illustrated as a scale-free network with the partner distribution *P* (*k*) ≈ 1*/k*^1.8^. This exponent implies a huge variation in connectivity and a core group that dominates the spread of the disease.

The significant variability in sexual risk behavior makes the concept of herd immunity more complex for mpox compared to diseases like influenza, where most individuals have a relatively equal likelihood of contracting and spreading the virus. In the homogeneous case, achieving a twofold decrease in the reproduction number requires immunization of half the general population. In the case of mpox, transmission is driven solely by individuals that are likely to have more than one unique sex partner in 2-3 week period, ie. the infectious period of mpox (Fig. 1 A). Thus, mpox herd immunity relies only on the immunity or sexual behavior change of a small subset of the population with many partners (6). At the same time, since high-risk activity symmetrically increases a person’s rate both of acquiring *and* transmitting the disease, the individuals that must be immunized for herd immunity are precisely the ones that will most rapidly contract the virus. Qualitatively, this logic predicts a rapidly increasing but short-lived mpox epidemic ending when the core group obtains herd immunity.

However, numbers from the actual 2022 epidemic challenge this prediction. Firstly, as seen in Fig. 1 B, The prevalence of mpox varied significantly across countries throughout the epidemic. Nevertheless, each country experienced a similar and synchronous decline in the reproduction number following the initial phase, which was characterized by superspreader events and high stochasticity due to the low number of cases. This indicates that an effective form of mitigation occurred simultaneously across countries in 2022. In this study, we pick up on the hypothesis by Zhang et al. (10) that the marked decrease in the serial interval observed during the epidemic (Fig. 4 A) may be explained by a shortened effective infectious period resulting from responsive behavior change (rather than an adaptation of the virus). We use our model to show that this phenomenon can alone explain the observed decline of the European epidemic.

The difficulty in quantifying the effect of natural herd immunity is due, in part, to the short infectious cycle of mpox (2-3 weeks (14)), which highlights the complexities in analyzing networks that change over time. In network theory, the *degree* of a node refers to the number of other nodes it is connected to. In the context of sexual networks, it is unclear how to apply this term - does a connection mean that you have had sex once ever, once recently, or that you have sex regularly? If we instead use the term *T-degree* to refer simply to a person’s number of unique partners in a time interval *T*, we can see that a person’s *T*-degree is not generally proportional to *T*. Most obviously, someone in a monogamous relationship may have both a one-week- and a ten-year-degree of 1. More pertinently, one cannot assume that someone with a 12-month degree of 12 has a 1-month degree of 1, let alone a 1-week degree of 1/4. This would only be true on the assumption that sex partners always have only one encounter, rather than a sexual relationship lasting some period of time. This is crucial for mpox, because a person’s infectiousness is determined by their 3-week degree, as illustrated in Fig. 1 A, while existing studies usually measure 1-year degrees.

Lacking data for the 2-week degree distribution among MSM, we approach the problem indirectly by building a self-organizing model in which agents build the network themselves by establishing connections according to a few simple rules. We then fit the model not only to the measured 1-year degree distribution but also the distribution of sexual encounter rate, probability of being in a steady partnership, and probability of a partnerships existing for a year: observations that provide hints as to the way in which partnerships form and dissolve over time. We call the model *Transient Attachment Network alGOrithm*, or TANGO.

Our behavioral data comes from the baseline survey of the Danish cohort study Project SEXUS (15). The survey included 62,675 individuals, selected to be nationally representative of gender, age, and geographic regions. This sample included 677 MSM (2.2% of male respondents), defined as those who reported at least one sexual encounter with another man in the previous year. We find that the 1-year degree distribution in this group follows a power law with exponent ≈ 1.8 (Fig. 2, B) in agreement with studies from the UK (16) and Netherlands (17). We assume that the observational data on sexual activity derived from Project SEXUS also apply to other European countries, including the fraction of MSM in a population and the partner distribution.

**Fig. 2.**
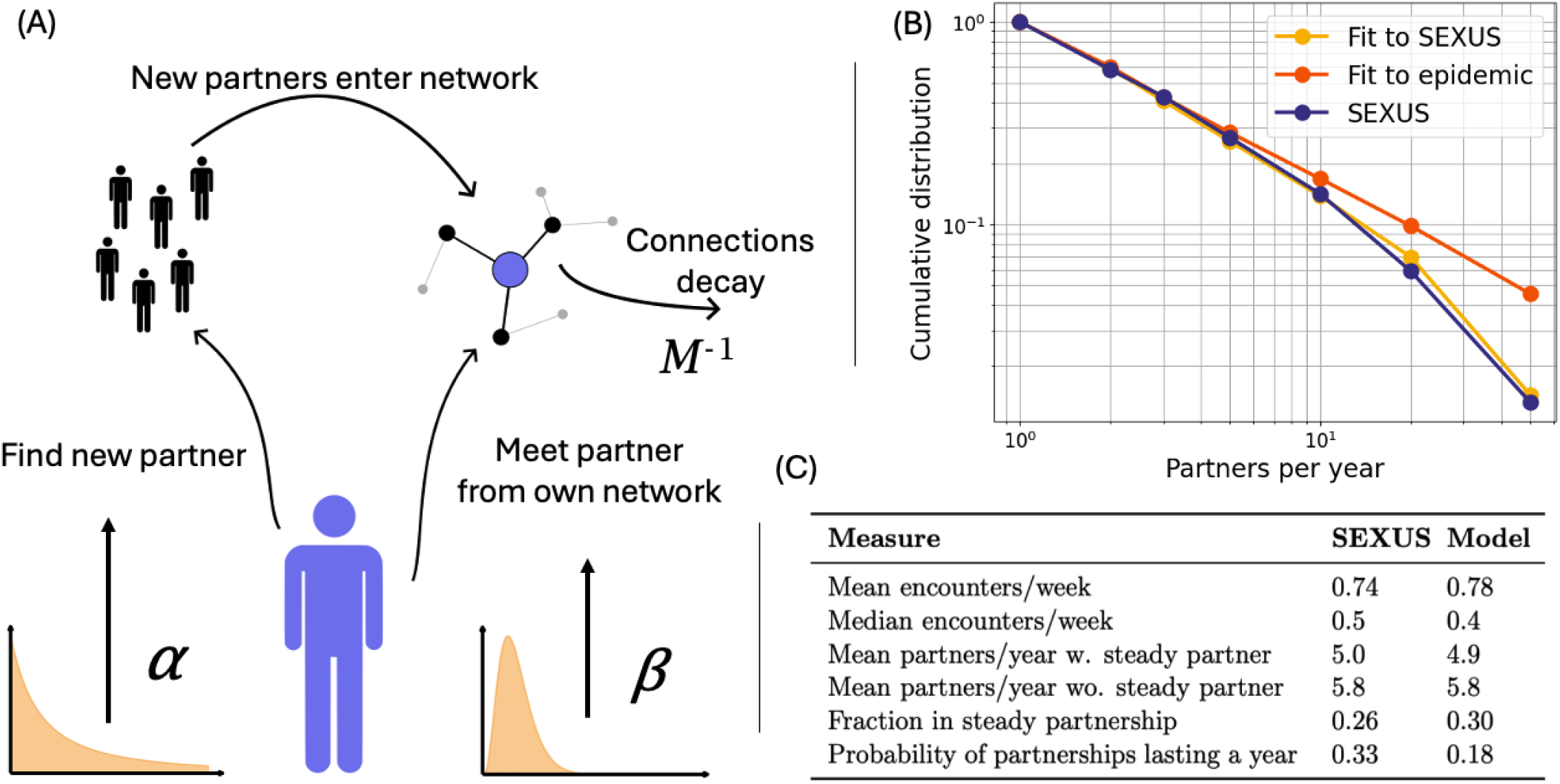
Model dynamics and data for pair formation in Danish MSM. **(A)** Cartoon of the dynamics in the TANGO model (*Transient Attachment Network alGOrithm*). Each day, an agent can either seek encounters with new partners, and establish new connections, or can connect to partners that he have already had sex with. An individual *i* is characterized by 1) the rate of seeking encounters with new partners (*α*_*i*_) and 2) the rate of seeking encounters with previous partners (*β*_*i*_). Connections decay at the rate 1*/M*, which thus limits the duration of sexual partnerships. **(B)** The parameters can be adjusted to reproduce the yearly degree distribution as it is reported in the Project SEXUS survey. After fitting to Project SEXUS, we fix 5 of 6 parameters, but use *b*_*u*_ to fit to epi curves and degree distribution of cases. A good fit is achieved with *b*_*u*_ = 250 partners/year, which produces the orange curve. **(C)** Comparison Project SEXUS survey data and the final model. Network simulations are run with a population of *N* = 50000 agents, and best-fit parameters are shown in the supplementary material.

## Results

### Fitting the model to survey data

We fit the TANGO model to reproduce multiple observations from the Project SEXUS data simultaneously, namely: the 1-year degree distribution, mean and median number of sex encounters per week, difference in yearly number of partners between those with and without a steady partner, and the dynamics of formation of steady partnerships. Fig. 2 shows agreement between the distribution of the number of partners during the last year observed in Project SEXUS (blue) and the model (yellow). The curve corresponds well to a power law with an exponent of 1.8 and a hard cutoff at 80 partners per year.

Fig. 2 C shows the agreement between Project SEXUS statistics and the model on other parameters. We have generally achieved a good fit, though partnerships are a bit too unlikely to last a year. The observables are selected to be descriptive of the dynamics of partnerships formation and dissolution. As the model can produce both one-time, casual (short-lived or low activity) and steady partnerships, it represents sexual network dynamics on both short and long timescales.

### Simulation of an mpox epidemic without mitigation

In addition to the degree distribution of the general MSM population, the epidemic model must reproduce the degree distribution of infected cases on both long and short timescales. Fig. 3 A compares the model and data from various countries on two timescales, 21 days and 90 days. Notice that there is less than a factor 4 difference in partner numbers between the two timescales, indicating concurrent partnerships of finite duration. In other words, some of the partnerships of the last 21 days likely also existed before that time window. Our model reproduces the activity distribution reasonably well on both time scales.

**Fig. 3.**
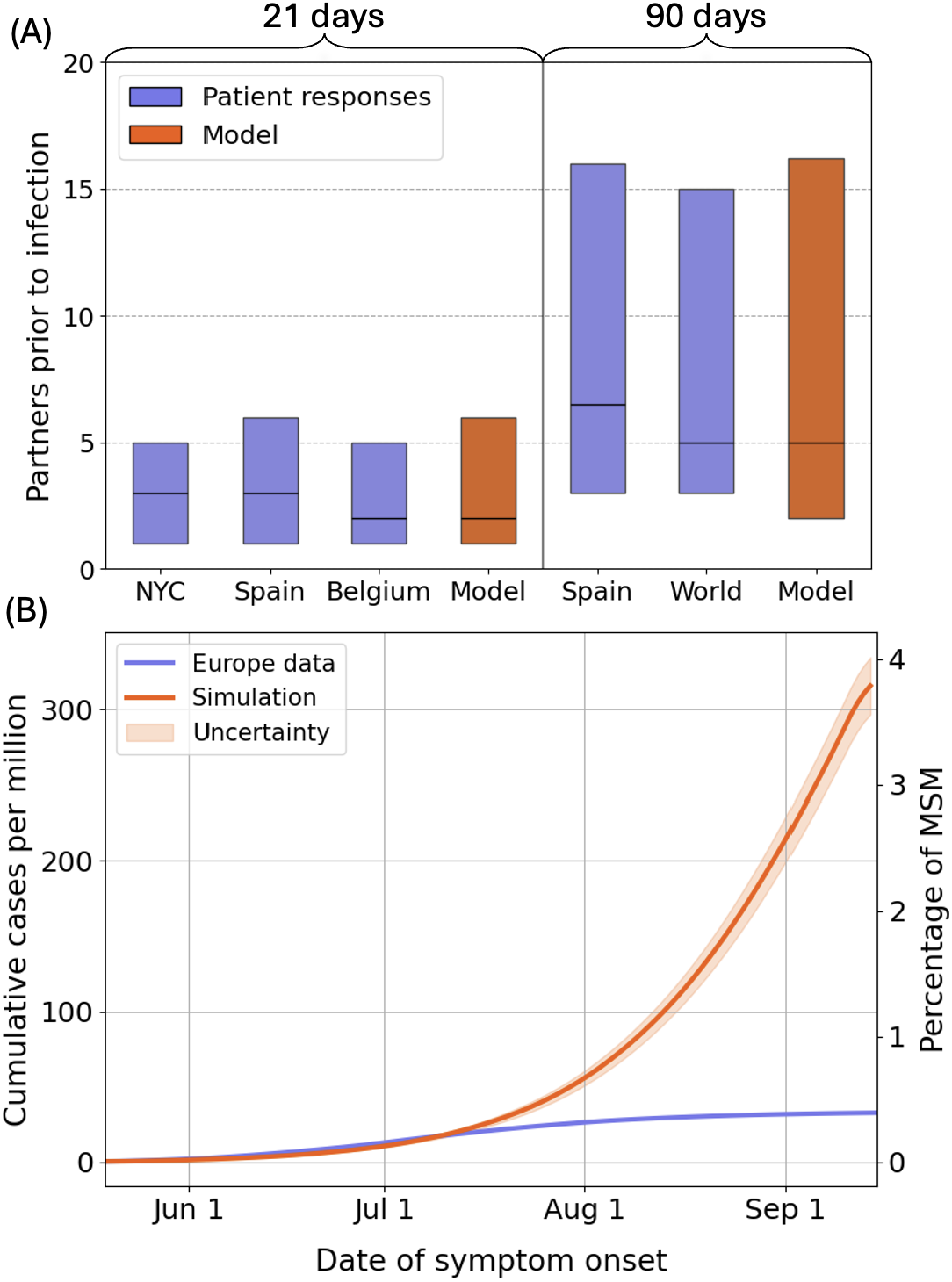
Model prediction of an mpox epidemic without mitigation. **(A)** Statistics of recent sexual activity in mpox cases in data (orange) and the model (purple), asking about the number of partners within both 21 days and 90 days before mpox diagnosis. In both model and observational data the median partner rate among infected persons is far greater than that of uninfected MSM (5 per 3 months vs. 2 per year). **(B)** The model predicts an epidemic that far exceeds the observed one, even before vaccination is initiated. Even though the model reproduces the activity patterns of cases well, it predicts a much higher threshold for herd immunity than possible with the total cases reported in Europe. The orange curve is a mean of 62 runs with a population *N*_*eff*_ = 100, 000, each starting with 6 seeded cases, picked randomly but with a probability weighted by *α*. Cumulatively, this corresponds to modeling the entire European MSM population. The filled orange area is the uncertainty from a single run divided by 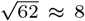, so it is our estimate for the uncertainty of the European cumulative curve.

Despite this agreement, the model predicts an epidemic drastically larger than that observed in any affected country in 2022, in the absence of any mitigation. In the model, exponential growth continues well into August, and only levels off visibly during September. Letting the epidemic run out (not shown in plot), we end at a mean final size of ≈ 900 infected per million. Assuming the actual European final size of 34 per million is subject to a 60% reporting rate (18), our model still predicts an outbreak 16 times greater. We conclude that the role of herd immunity was negligible by the time the 2022 mpox epidemic peaked.

### Simulation of an mpox epidemic with mitigation by responsive behavior change

Fig. 4 A shows the decrease in serial interval documented in (10) along with a fit to a sigmoid function

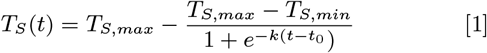

with *T*_*S,max*_ = 14 days, *T*_*S,max*_ = 6.5 days, the characteristic rate *k* fitted to 2.45/month and the day of steepest descent, *t*_0_, fitted to June 6.

**Fig. 4.**
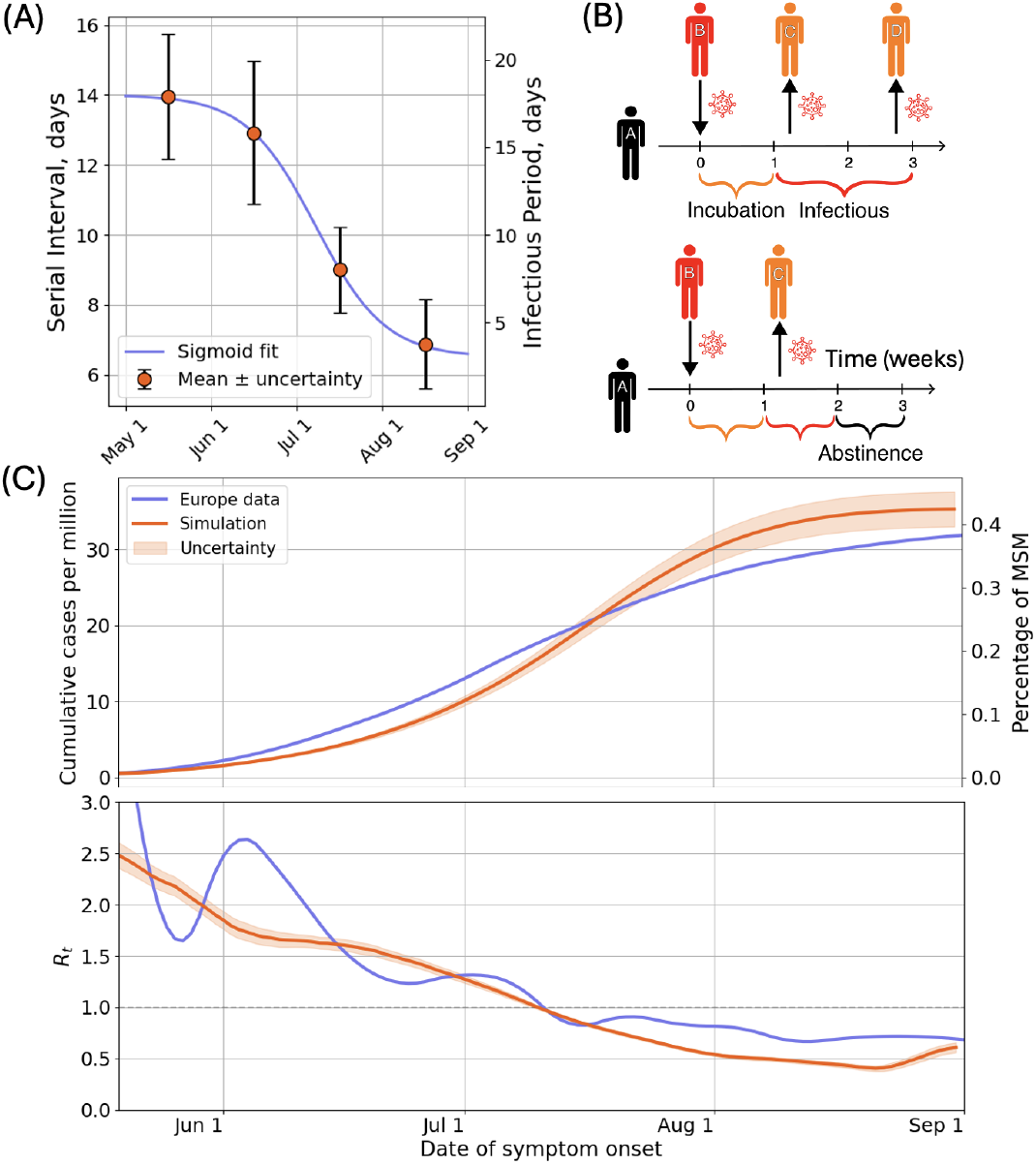
Introducing responsive behavioral change. **(A)** Observed shortened serial interval during the epidemic (10) is consistent with responsive behavior change. The blue line shows a fit used to parametrize the time dependence of the effective infectious period. The right *y*-axis shows the effective infectious period according to eqn. 2. **(B)** Illustration of how behavior changes lead to a decrease in the serial interval and the number of secondary cases. In the top diagram, the measured generation time (which approximates the serial interval) is 2 weeks, as this is the mean between 1 and 3 weeks. In the bottom diagram, abstinence in response to symptoms means that only the early transmission occurs, and the average serial interval will be only 1 week. **(C)** With this behavior modification the model tracks the epidemic curve in Europe. As a consequence, the effective reproductive number also tracks the observation. As in Fig. 3 B, the orange curve corresponds to 62 runs of *N*_*eff*_ = 100, 000. Clearly, the uncertainty from a single run is considerable, which we argue in supplementary material is in accordance with reality.

Fig. 4 B illustrates how responsive behavioral change may explain a shorter serial interval. Increased awareness about the symptoms and seriousness of mpox leads people to respond more readily to symptoms by going into abstinence. The mean serial interval is an average over transmission events, some of which happen early in the biological infectious period, while others happen late. However, responsive behavior change will lead to fewer late transmissions, and thus a reduction of the mean serial interval.

If we assume a constant rate of infection from the end of the latent period *T*_*E*_ until the end of the effective infectious period, *T*_*I*_ (due to recovery or abstinence), then the serial interval *T*_*S*_ will be given by

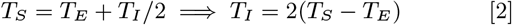

Thus, we can translate the measured serial intervals into decreasing infectious periods, which we can insert into our model, and the result is seen by reading the right *y*-axis on Fig. 4 A. Notice that the inferred decrease in the effective infectious period is by a factor of 6; from 18 days to about 3.

If we repeat the simulation with a gradual decrease in effective infectious period as in Fig. 4 A, we find that this responsive behavior change is sufficient to explain the decline of the European mpox epidemic (Fig. 4 C): In fact, the reduction in the effective reproduction number *R*_*t*_ in the latter half of the epidemic is too steep, with the epidemic almost dying out before September 1st. This latter discrepancy may be due to different factors: 1) uncertainty in the August measurement from Zhang et al., which is based on less than half as many datapoints (15) as the previous months, 2) that our model assumes that every individual behaves equally responsively and timely while it may be that there exists a less responsive sub-population that is less inclined to change behavior, and 3) be due to the uncertainty in determining the latent period. In equation 2, once *T*_*S*_ ≈ *T*_*E*_, small uncertainties in *T*_*E*_ have a large relative effect on the predicted *T*_*I*_. In any case, the model demonstrates that the responsive behavior change indicated is sufficient to explain the remarkably early decline of the epidemic.

### Synchronous reaction across European countries

As we saw in Fig. 1, the gradual decrease in reproduction number was remarkably synchronous across European countries. The data appear to be consistent with a model in which MSM across European countries are reacting in lockstep, irrespective of prevalence in the individual countries. It may be that the ramping up of mpox information campaigns led to similar degrees of responsive behavior change across different countries at the same time, perhaps owing to appeals and guidelines from international agencies (4)(19).

To test this hypothesis, we repeat the simulation with time-dependent *T*_*I*_ as in Fig. 4, but with introduction dates mimicking the known introduction dates of mpox in different countries. Specifically, we introduced mpox cases corresponding to 1 per million population (roughly 1 per 10,000 MSM) at the respective dates when countries reached this number of cumulative cases. For each country, the resulting size of the final epidemic is then compared to the observed case count in Fig. 5.

**Fig. 5.**
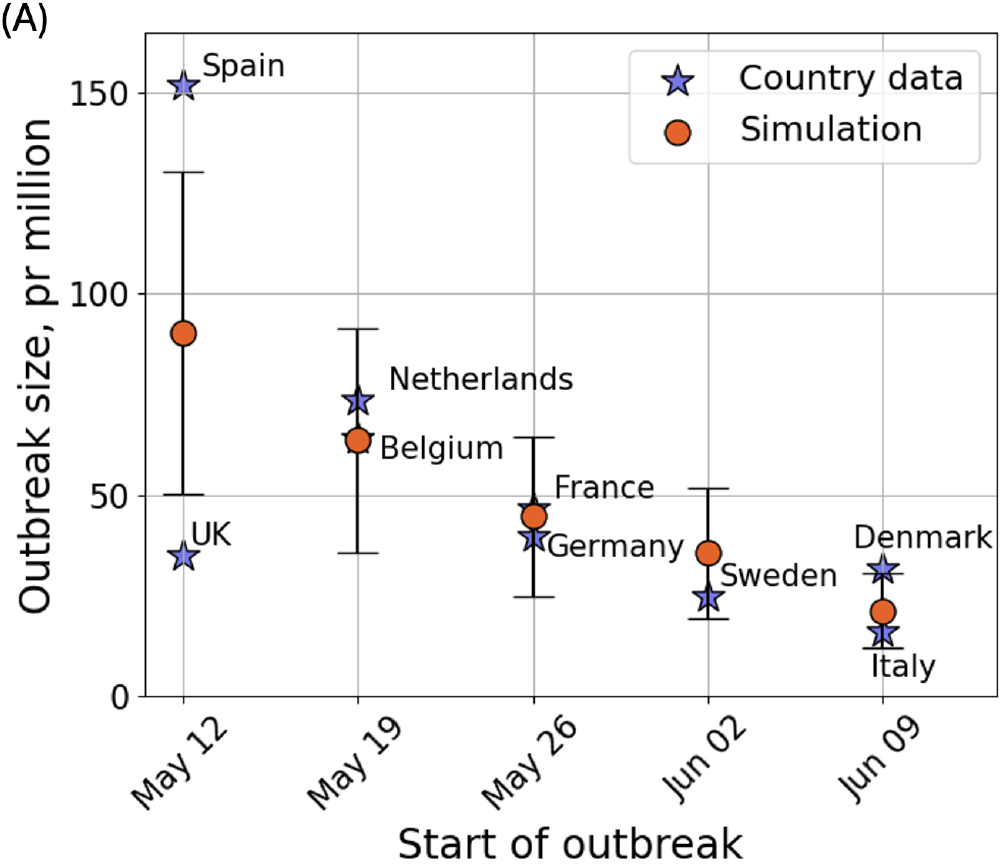
Outbreak size depends on the time of mpox introduction. The “start of the outbreak” in a country is defined as the first day where the cumulative cases surpass 1 per million. Countries whose epidemics started late recorded smaller outbreaks. This indicates that awareness and resulting responsive behavior change traveled faster than the epidemic. Running model simulations with different introduction dates but keeping behavior change from Fig. 4 A, reproduce the observed country systematics. The dots represent means of 50 runs, and the error bars are simply the standard deviation of the results.

Apart from the UK, the country data show a clear decrease over time in outbreak size, indicating a clear advantage to having the virus introduced late. This trend is reproduced well by the model. The unusually large outbreak in Spain is probably due to the fact that the biggest superspreader events - Maspalomas Gay Pride and the outbreak in a Madrid Sauna (3) - happened there. At the same time, the conspicuous outlier of the UK may indicate that this country reacted more strongly against the epidemic than other countries, as evidenced by observations of a decline in other STIs that was not seen in the Netherlands and Belgium (20).

## Discussion

We designed a model of sexual pair formation in an MSM population and fitted it to several bulk observations from Project SEXUS (Fig. 2), a large Danish study that involved 677 randomly sampled MSM. When we then used the model to simulate the mpox epidemic on the network, it also reproduced the number of partners of mpox cases on both the short (21 days) and long (90 days) term (Fig. 3 A), indicating that our model is a good representation of the core group dynamics governing mpox propagation.

With the model thus verified against data, we show that Europe was nowhere near herd immunity when the epidemic peaked (Fig. 3 B) and that the outbreak would have been nearly 20 times larger without mitigation. Based on a simple assumption of constant infection rate during the mpox infectious period (eq. 2), we use data for the decrease in mpox serial interval over time to infer that the effective infectious period decreased from around 18 days in May to around 3 days in September (Fig. 4 A). Our model shows that this was more than enough to explain the decline of the European epidemic (Fig. 4 C). Finally, we use the model to show that the country-level data is consistent with a synchronous responsive behavior change across European countries (Fig. 5).

We believe ours is the first network model of mpox to be fitted using data for reported activity of mpox cases. One previous modeling study (10) explicitly checks their results against this data and finds a substantial discrepancy, stating that this may be explained by social desirability bias. While social desirability bias could plausibly play a role – patients who are handed a survey at the same time as they receive a serious diagnosis may be hesitant to provide honest answers about risky sexual behaviors – the consistency of the data across different countries (Fig. 3 A) may suggest that the data are reliable.

Our model’s agreement in terms of activity of mpox cases on both the short and long timescale stems from the fact that our model represents sexual relationships of finite duration overlapping in time. The problem of how to represent *concurrent* partnerships was addressed in several studies following a seminal article by Kretzschmar and Morris (21), but has largely been ignored in the context of mpox. We believe that the short infectious period of mpox makes concurrency considerations inevitable in this context.

Though the method of using *b*_*u*_ as a fitting parameter is a highly speculative best attempt at representing sexual risk behavior among European MSM, it does not weaken the assertion that herd immunity was not the cause of the epidemic’s decline. On the contrary, if we had held *b*_*u*_ fixed to the value suggested by Project SEXUS (Fig. 2 B), this would have corresponded to a more homogeneous network and thereby an even higher threshold for herd immunity, assuming *p* had been adjusted to achieve the correct reproduction number.

A core assumption in our work is that the observed decrease in serial interval for mpox in (10) can be attributed to responsive behavior change, rather than some change in the biological properties of the adapting virus. We assume that, over time, mpox-infected individuals more readily go into abstinence in response to symptoms, shortening the effective infectious period. This may reflect an increased knowledge about the symptoms and potential severity of the disease, as well as increasing contact-tracing efforts. The authors in (10) suggested the same interpretation, but inferred a far less dramatic decrease in effective infectious period (about 20%). This marked difference is likely because they allow for a preventive behavior change towards less risk behavior in the general mpox-free MSM population, in addition to responsive behavior change among those with mpox infection. They model both changes simultaneously to achieve the best fit to the epidemic trajectories. However, upon reviewing the literature on the prevalence of other sexually transmitted infections (STIs) among the core group during the 2022 mpox outbreak, we found no evidence of a decrease in sexual risk behavior in the general MSM population. Studies from Belgium (12) and the Netherlands (13) showed no change in the prevalence of endemic STIs among HIV-PrEP users during the epidemic. Similarly, the median number of partners in the previous 3 months among mpox-free participants in a clinical trial involving French PrEP users was unchanged between May-July and July-September (22). Accordingly, we took a different approach than (10), committing to the reduction of *T*_*I*_ that a simple model would predict from the *T*_*S*_ time series (Fig. 4 A, eqs. 1, 2) and then accepting an imperfect fit to the epi curves (Fig. 4 C). The goal, then, is not to make a perfect replication of the epidemic with an imperfect model but to pinpoint the most substantial cause of the epidemic’s decline.

The synchronicity with which the reproduction number dropped in different European countries in 2022 motivated us to compare our results to the epi curve for Europe as a whole, rather than that of a single country. This approach largely avoids complications stemming from import and export of cases. Clearly, even though it is evident that the reproduction number decreased synchronously across Europe, we cannot say for certain that the cause was the same everywhere. Indeed, a significant problem with our analysis is that the observation of decreasing serial interval comes from the UK (10) - precisely the country that is least representative of the general epidemic trend in Europe (Fig. 5). We do not know of any similarly detailed measurements of the changing serial interval, and the UK measurements of the serial interval are the most commonly cited. It is possible that the earlier peak in the UK is mainly due to preventive behavior change, and that the level of responsive behavior change among UK MSM was representative of Europe, as we assume, but we cannot know.

Despite the mpox epidemic effectively fading in the latter half of 2022, the clade IIb virus is now endemic and spreading globally. This was in part documented in a study that found mpox DNA in discarded condoms in Asian brothels and parks during 2023 (23). Surprisingly, in 2024 Australia even saw an outbreak among MSM larger than the one of 2022, with about 1300 cases (5). Meanwhile, European countries have reported many introductions in the years following 2022, but no large outbreaks (5).

As vigilance against mpox has probably worn off due to vanishing risk, a country’s future susceptibility to mpox is likely determined by its vaccine coverage among the core group. It is therefore remarkable that in Australia, 50% of MSM report having received at least one vaccine dose (24), while only about 10% of Danish MSM have been vaccinated (data from Statens Serum Institut). Heterogeneity offers a possible explanation for how Australia can have both a larger vaccination coverage and a higher susceptibility. While the European vaccination strategy was to target the core group of high-risk MSM, using criteria such as the recent history of multiple partners, participation in group sex or receiving HIV-PrEP (25)(26), official websites of Australian health authorities indicate that a less targeted approach was taken there (27)(28), offering the vaccine more broadly to the entire MSM population. In Denmark, the vaccine is offered only on the condition of either HIV positivity, HIV-PrEP use, or eligibility for PrEP (having multiple partners) (26). With around 4,000 Danish PrEP users (8% of MSM) (29), a large majority of the Danish core group has likely received at least one dose. In a Danish HIV cohort, 77% of participants with more than 5 partners in 12 weeks had received the vaccine at a follow-up 6 months after the epidemic (30). We could not find similar data for Australia.

The analysis and model presented here provide a framework that can aid in designing effective mitigation strategies for STIs in the future and making reliable predictions on their effects. As yet another mpox clade (clade Ib) now spreads human-to-human in central Africa, the targeted vaccination efforts initiated in 2022 are likely shielding the European MSM community from new outbreaks. The 2024 spread of mpox among also women and children in Congo could be analyzed through a variation of our model that includes the heterogeneous sexual activity of female sex workers and their male customers combined with a secondary spread between mother and child.

## Materials and Methods

### The TANGO model

The idea of the model is illustrated in figure 2 (A), with a detailed description in the supplementary material. It is executed in time-steps Δ*t*, where each agent may attempt to find a sex partner. In a time-step, an agent *i* will, with probability Δ*t* · *α*_*i*_, seek a *first-time encounter* with someone he has not had sex with before, and with probability Δ*t* · *β*_*i*_ seek a *network encounter* with someone he knows from previous encounters. If *i* has a first-time encounter with *j*, an edge is drawn between *i* and *j* with weight *σ*_*i,j*_ = 1. If later *i* and *j* have a network encounter, *σ*_*i,j*_ increases by 1 for every encounter. The rate of encounters between *i* and *j* is proportional to *σ*_*i,j*_, reflecting a stronger connection. This implies positive feedback that enables long-time partnerships. At the same time, connections decay at rate *σ*_*i,j*_ · Δ*t/M*, where *M* is a “memory” parameter assumed to be the same for all individuals. The positive feedback is thus balanced by two limiting effects: (1) Connections decay at a rate proportional to their weight and (2) the rate of encounters between *i* and *j* is limited by their rates of seeking network encounters - *β*_*i*_ and *β*_*j*_ - which are constant person-specific parameters.

Executing network encounters is not trivial, because an encounter between *i* and *j* requires two conditions to be satisfied simultaneously: 1) both *i* and *j* are seeking a network encounter in this time-step and 2) neither *i* nor *j* have a network-based meeting with someone else in this time-step. To achieve this, network encounters are executed by picking out *edges*, rather than agents, with a probability proportional to their weight. See supplementary material for a detailed explanation.

When an agent is infected, his infection cycle follows the SEIR (susceptible, exposed, infectious, recovered) form. While the incubation period has been measured at around 8 days (31), substantial presymptomatic shedding and infection have been documented (32) (33), and measurements show a median interval of 5 days from exposure to viral shedding (32). Thus, we set the mean latent period *T*_*E*_ to 5 days. We model all state transitions as a two-step Poisson process, meaning that the transition time from state X follows an Erlang distribution with mean *T*_*X*_ and shape factor 2.

We let the *I*-state denote effective infectiousness, such that an individual’s choice of entering abstinence while still infectious is implemented as transitioning to the recovered state. For this reason, we allow *T*_*I*_ to change over time as a reflection of responsive behavioral change. *T*_*I*_ (*t*) is given by eqs. 1 and 2.

The model goes through two rounds of adjustments of parameters for, respectively, the network dynamics and the disease transmission dynamics. Thus, the parameters describing partnership formation are fitted to Project SEXUS first. Subsequently, the secondary attack rate *p* and the cut-off *b*_*u*_ to the number of new partners per year *α*_*i*_, are fitted to yield a good representation of the epidemic in terms of the activity of cases and the reproduction number.

### First round of fitting: Project SEXUS

The network model has six parameters: Upper and lower bound to the *α*-distribution, *b*_*u*_ and *b*_*l*_, power-law exponent to *α*-distribution, *γ*, mean rate of seeking new encounters *β*_0_, shape parameter to *β*-distribution *k*_*β*_ and connection decay-rate *M*. These parameters are jointly adjusted to achieve a good fit to the 1-year degree distribution (which will differ slightly from the *α*-distribution), as well as the observables shown in Fig. 2 C). The parameters *b*_*u*_ and *b*_*l*_ are necessary due to the fact that the power-law exponent of the 1-year degree distribution is *<* 2 (7) (17).

Note that steady partnerships are not explicitly built into the model. Instead, we make a heuristic definition of steady partnerships within the model, which allows them to arise based on the model rules. Suppose agents *i* and *j* are connected with weight *σ*_*i,j*_. Now, the partnership is a steady partnership if, *σ*_*i,j*_ *>*∑_*k* ≠ *j*_ *σ*_*i,k*_, *σ*_*i,j*_ *>*∑_*k*≠ *i*_ *σ*_*j,k*_ and *σ*_*i,j*_ *>* 1. Roughly speaking, this means that two people are steady partners if they each currently direct more than half of their sexual activity to the other person, and if they have had sex more than once within a prior period *M*. This allows each person to have no more than one steady partner, similar to the formulation in Project SEXUS, which did not allow respondents to report multiple steady partners.

### Second round of fitting: Epidemic trajectory and degree distribution for infected individuals

Though we have already fitted the six network parameters, we leave *b*_*u*_ as a free parameter to achieve a good fit to the degree distributions of infected cases (Fig. 3 A) and the growth rate in the early epidemic. The reason for this is as follows. Since activity at the tail of the distribution - where the core group is found - effectively drives the epidemic, the value of the cut-off is a crucial parameter. Incidentally, it is also a difficult parameter to estimate, because the number of people constituting the tail is small, and therefore an enormous sample size is required to represent them in a survey. Comparing studies of the general population with ones selected for high-risk individuals, e.g. questionnaires given to STI patients, yields markedly different estimates of this cutoff (16). In light of this, it seems likely that high-risk behavior correlates negatively with participation in general population surveys, and that such studies will systematically underrepresent the tail of the sexual degree distribution. As the baseline survey of Project SEXUS represents such a study, we allow *b*_*u*_ to be a free parameter that we can use to fit the course of the epidemic, while keeping all other network parameters fixed to the values obtained in fitting to behavioral data.

As the mean latent period *T*_*E*_ was estimated from (32) and *T*_*I*_ (*t*) is estimated from *T*_*S*_(*t*) (Fig. 4 A), this leaves just one biological parameter, the secondary attack rate, *p*, to be adjusted together with *b*_*u*_.

All epidemic simulations in this paper use an effective population size of *N*_*eff*_ = 10^5^, corresponding to twice the Danish population of MSM. Please see supplementary material for the explanation of *N*_*eff*_.

## Data Availability

The survey data used in this study are not publicly available due to confidentiality agreements and participant privacy protections. Simulated data supporting the findings of this study are available upon request. All code used for data processing and analysis will be made publicly available on GitHub prior to publication.

## ACKNOWLEDGMENTS

The authors would like to thank Viggo Andreasen for his theoretical contribution. We would also like to thank Maria Wessman and Anders Koch for data on mpox vaccines in Denmark and Mikael Andersson for the Project SEXUS data. This research was funded by the Danish National Research Foundation (grant no. DNRF170).

