## Supplementary material for "Relationship dynamics and behavioral adaptations in the control of the 2022 mpox epidemic"

### 1. Supplementary material

**A. The TANGO model.** The TANGO model (*Transient Attachment Network alGOrithm*) combines well-mixed and network dynamics (Fig. 2 A) to generate relationships of varying length. An iteration of the pair-formation algorithm has three phases:

- *Choice phase*: Agents choose which encounters to seek in this iteration.
- *Pairing phase*: Agents are paired according to these choices.
- *Decay phase*: Connections decay stochastically.

We distinguish between two types of encounters: first-time encounters and network encounters, with the latter referring to encounters between two agents who have a connection due to a previous sexual encounter. First-time and network encounters are governed by the rates $\alpha_i$ and $\beta_i$, respectively, with $i$ an index referring to the agent.

***A.1. The choice phase.*** In the choice phase, agents decide whether they will seek first-time and network encounters today. To determine whether agent $i$ is seeking first-time encounters in an iteration, draw a number from a Poisson distribution with mean $n_{f,i} = \mathrm{d}t \cdot \alpha_i$. Then, $n_{f,i}$ is the number of times that $i$ will seek a a first-time encounter in this iteration. If $dt = 1$ day, as in our simulations, $n_f$ will typically be 0, occasionally 1, for anyone with $\alpha_i \ll 365$/year, which applies to the vast majority. For a very high value of $b_u$, some people will occasionally have 2 new partners in one day.

At a given time, each member is characterized by a number $s_i$ which is either 1 or 0, 1 indicating that $i$ is seeking a network encounter and 0 meaning that he is not. In the choice phase, an agent $i$ who is not currently seeking will become seeking (switch $s_i$ from 0 to 1) with probability $\mathrm{d}t \cdot \beta_i$. He will remain seeking until he has a network encounter.

***A.2. The pairing phase.*** Network encounters are executed before first-time encounters, such that first-time partners of this iteration are not candidates for network encounters in the same iteration.

For network-encounters, make a copy of the network and prune it such that an edge $(i, j)$ remains in the copied network only if $s_i = s_j = 1$, ie. if both parties are seeking a network encounter in this iteration. Now, list all edges in the pruned network and call this list $L$. $L$ is then a list of 2-tuples $(i, j)$. Pick a random edge from $L$, weighting the probability of picking edge $(i, j)$ proportionally to $\sigma_{i,j}$. This is the positive feedback underlying the model: encounters are more likely to happen between strongly connected agents.

After picking $(i, j)$, make the substitutions $\sigma_{i,j} \to \sigma_{i,j} + 1$, $s_i \to 0$ and $s_j \to 0$ (connection is strengthened and agents are no longer seeking network encounters). Now prune the network again by removing from $L$ all edges that are connected to either $i$ or $j$, ie. all connections of the form $(x, i), (i, x), (x, j)$ or $(j, x)$ for any $x \neq i$ or $j$. Repeat this process until the pruned network is empty. This is the most computationally intensive part of the code.

For first-time encounters, make a list containing every agent who is seeking a first-time encounter in this iteration. Let agent $i$ appear $n_{f,i}$ times in the list, where $n_{f,i}$ is the number of times agent $i$ is seeking first-time encounters, determined in the choice phase. Next, pick two random elements (agents) on this list. If the two elements are $i$ and $j$, draw an edge between $i$ and $j$ with strength $\sigma_{i,j} = 1$, and delete the two chosen elements from the list. Continue until there is only one or zero unique agents in the list.

Notice that the model is structured such that everyone seeking a first-time encounter in this iteration will get it (sometimes except one person), while not everyone seeking a network encounter will get it. If agent $i$ is seeking a network encounter ($s_i = 1$), he will still only get it if at least one of his connections is also seeking ($s_j = 1$) AND that person does not have a network encounter with someone else first.

This is why we have chosen to use the boolean $s_i$, which stays at 1 if $i$ does not have a network encounter in this iteration. If the decision of whether to seek a network encounter was made independently each day, the probability that $i$ is seeking AND that one of his partners is seeking would go as $\mathrm{d}t^2$, and parameters would have to change substantially with changing $\mathrm{d}t$ to preserve behavior.

***A.3. The decay phase.*** The decay phase is simple: for each edge $(i, j)$ with strength $\sigma_{i,j}$, draw $\sigma_{i,j}$ random numbers. For each random number, evaluate whether it is below $\mathrm{d}t/M$. If so, substitute $\sigma_{i,j} \to \sigma_{i,j} - 1$. In this way, connections decay at a rate proportional to their strength, preventing connection strengths from diverging.

***A.4. Network parameters.*** As explained in the main text, the model goes through two rounds of fitting, first fitting only the relationship dynamics to the data from Project SEXUS, and subsequently fitting to the measured serial interval, reported number of partners among mpox cases and the early epidemic growth rate. The best fit to Project SEXUS is obtained with an upper bound $b_u = 70$ to the $\alpha$-distribution. As argued in the main text, we believe the true number is higher, and thus we keep this crucial parameter free for the second round of fitting.

Note that the blue and yellow curves in Fig. 2 B correspond to a power law with upper bound 80 and exponent 1.8, not 70 and 1.55 as in table 1. This is because the latter variables describe the distribution of the yearly rate of seeking *new partners*. However, this is not precisely the same as the number of *any partners* in the last year, since some of these partnerships may have started more than one year ago (eg. a steady partner). It turns out that the parameters in table 1 are the ones that yield the correct power law for the number of partners in the last year.

| Name | Value | Interpretation | Source |
|---|---|---|---|
| $\beta_0$ | 0.2/day | Mean rate of seeking network encounters | Fitted to SEXUS |
| $k_\beta$ | 2 | Shape parameter for gamma distribution of $\beta$ | Fitted to SEXUS |
| $\gamma$ | 1.55 | Power law exponent on $\alpha$-distribution | Fitted to SEXUS |
| $b_l$ | 0.15/yr | Lower bound on $\alpha$-distribution | Fitted to SEXUS |
| $M$ | 28 days | Timescale of connection decay ("memory") | Fitted to SEXUS |
| $b_u$ (first fit) | 70/yr | Upper bound on $\alpha$ distribution | Fitted to SEXUS |
| $b_u$ (second fit) | 300/yr | Upper bound on $\alpha$-distribution | Fitted to epidemic |

**Table 1. Network Parameters**

| Name | Value | Interpretation | Source |
|---|---|---|---|
| $T_E$ | 5 days | Mean incubation period | (1) |
| $T_{I,0}$ | 18 | Mean effective infectious period, prior to behavior change | (2) and eq. 2 |
| $p$ | 0.48 | Secondary attack rate | Fitted to epidemic |
| $k_T$ | 2 | Shape parameter for Erlang time-distributions | (3) |
| $p_{vac}$ | 0.13 | Probability of protection from smallpox vaccination | (4) (5) |

**Table 2. Epidemic Parameters**

Our dataset includes everyone in the Project SEXUS baseline survey who reported at least one male sex partner in the last year. As our sexual network model is stochastic and self-organizing, there is no guarantee that every agent has any partners in a given year. Thus, if the simulated population has size $N$, there will in a given iteration be a smaller subpopulation $N_{eff}$ of people who actually had a partner in the last year. With the SEXUS-fitted parameters, about 42% of agents do not have sex in a given year. The measures in Fig. 2 C are all calculated after masking out these 42% inactive members. This fraction does not change substantially after increasing $b_u$ to 250. Thus, to achieve a population of $N_{eff} = 100,000$ requires a simulated population of $N = 170,000$.

While it is natural that the population of people who *might* have a male sex partner in a given year is larger than the population which actually has it, it is probably not realistic that the latter is 42% smaller than the former. For instance, in Project SEXUS, 12% of those who identify as homosexual have not had sex in the last year (6). This is unsatisfying, but not critical, since agents that do nothing have no effect on network dynamics.

**B. Test against well-mixed heterogeneous pairing.** In this section we present a brief analysis of how the predictions of our model differ qualitatively from those of a simpler model assuming well-mixed heterogeneous pairing but no network structure and no repeated encounters.

Consider a population divided into $n$ activity groups, where group $i$ has contact rate $c_i$, meaning the rate of picking a random partner from the population. The probability that your partner belongs to group $j$ is then proportional to $c_j$. With an infinite number of activity groups with power-law distributed concentrations of people, this corresponds exactly to the "first-time encounter" part of our model.

In such a model, the force of infection $\Lambda$ will be

$$\Lambda = \sum_i \frac{c_i I_i}{\sum_j c_j N_j} \cdot p = \zeta \sum_i c_i I_i, \qquad \text{[S1]}$$

$$\zeta \equiv \frac{p}{\sum_j c_j N_j}, \qquad \text{[S2]}$$

where $c_i, I_i$, and $N_i$ are the contact rate, number of infected, and total number of members of group $i$. Assuming exponentially distributed recovery time, and neglecting the latent period, the epidemic dynamics within group $i$ follow

$$\dot{S}_i = -c_i S_i \Lambda \quad \text{[S3]}$$

$$\dot{I}_i = c_i S_i \Lambda - \nu I_i \quad \text{[S4]}$$

where $\nu$ is the recovery rate. Now, using a trick from (7), we can ask about the derivative of $S_i$ wrt. the number of susceptibles in a certain other activity group, say $S_1$:

$$\frac{\mathrm{d}S_i}{\mathrm{d}S_1} = \frac{\dot{S}_i}{\dot{S}_1} = \frac{c_i S_i}{c_1 S_1} \implies \frac{1}{S_i} dS_i = \frac{c_i}{c_1}\frac{1}{c_1} dS_1 \quad \text{[S5]}$$

Integrating from 0 to $t$:

$$\log S_i(t) - \log S_i(0) = \frac{c_i}{c_1}\left(\log S_1(t) - \log S_1(0)\right) \implies \quad \text{[S6]}$$

$$\frac{S_i(t)}{S_i(0)} = \left(\frac{S_1(t)}{S_1(0)}\right)^{c_i/c_1} \implies \quad \text{[S7]}$$

$$S_i(t) = S_i(0) \cdot \left(\frac{S_1(t)}{S_1(0)}\right)^{c_i/c_1} = N_i \cdot \left(\frac{S_1(t)}{N_1}\right)^{c_i/c1} \quad \text{[S8]}$$

If we define $s_i(t) = S_i(t)/N_i$, this becomes

$$s_i(t) = s_1(t)^{c_i/c_1} \quad \text{[S9]}$$

Next, if we take $c_i$ to be continuously distributed, we can instead write

$$s(c,t) = s(c = c_1, t)^{c/c_1} \quad \text{[S10]}$$

Now, for $t \to \infty$ (at the end of an epidemic that has ended by natural herd immunity), $s(k,\infty) + r(k,\infty) = 1$, so

$$r(c,\infty) = 1 - s(c,\infty) = 1 - s(c = c_1,\infty)^{c/c_1} = 1 - (1 - r(1,\infty))^{c/c_1} \quad \text{[S11]}$$

This gives us a distribution that we can compare to our simulation. Let us choose $c_1 = 1$ /year. The probability $r(1,\infty)$ then depends on $\nu$ and $p$, but let us keep it as a free parameter. Then, setting $r(1,\infty) = 0.015$ or $r(1,\infty) = 0.025$, respectively, we obtain the curves in Fig. 1.

We see that $r(1,\infty) = 0.15$ gives a good fit at high $\alpha$, while $r(1,\infty) = 0.25$ gives a good fit at low $\alpha$. None of them are a good fit to the entire curve however. As a result of this difference, we find that it is not possible for this simpler model to reproduce the degree distribution for mpox cases on both timescales. Fig. 2 shows results for running our model with no memory, ie. with only first-time encounters, which is precisely the system described by the equations above. Using the same parameters, the model gives $R_0 = 2.0$, roughly equivalent to the early epidemic, and accordingly, the 21-day degree fits fairly well with data. However, the 90-day degree gets significantly too high, as one would expect - in this case it should be 4 times higher, up to rounding effects.

This is expected: The model that is described by eq. S11 is one in which, for instance, reporting 5 partners per year means having just 5 sex encounters in a year. Clearly, this puts you at very low risk of acquiring mpox, because even if one of your partners gets infected, you still need to . Since our model allows for repeated encounters between partners, it increases the risk for people with relatively few partners. The difference is less pronounced for individuals with high $\alpha$, because their sexual activity is more dominated by first-time encounters, and for them repeated encounters make a smaller difference, in relative terms.

**C. A noisy epidemic.** As we noted under Fig. 4, there is a substantial uncertainty to the size of an epidemic when seeding 6 cases in a population of $N_{eff} = 100,000$ MSM. The 6 seeds are chosen with a probability weighted by their rate of seeking new partners $\alpha$. This large degree of noise is an interesting feature of the heterogeneity of the network. Despite $R_0 > 2$, many seeds will not produce an exponentially growing transmission chain, or will produce one that stays below exponential for several infectious cycles, giving responsive behavior change the chance to kick in in the meantime. As a result, founder effects introduce substantial uncertainty in the final size of the epidemic.

Qualitatively, this effect can be observed in epidemic data (Fig. 3. Countries that had few importations, in absolute terms – mainly small countries – displayed very noisy, irregular epidemic curves very unlike the classical epidemic bell curve 3. This indicates that these founder effects also introduced uncertainty in the actual epidemic, and only larger countries with many importations from the early superspreader events in Spain converged stably on a similar growth rate 1.

1. I Brosius, et al., Presymptomatic viral shedding in high-risk mpox contacts: A prospective cohort study. *J. Med. Virol.* **95** (2023).
2. XS Zhang, et al., Transmission dynamics and effect of control measures on the 2022 outbreak of mpox among gay, bisexual, and other men who have sex with men in england: a mathematical modelling study. *The Lancet Infect. Dis.* **24**, 65–74 (2024).

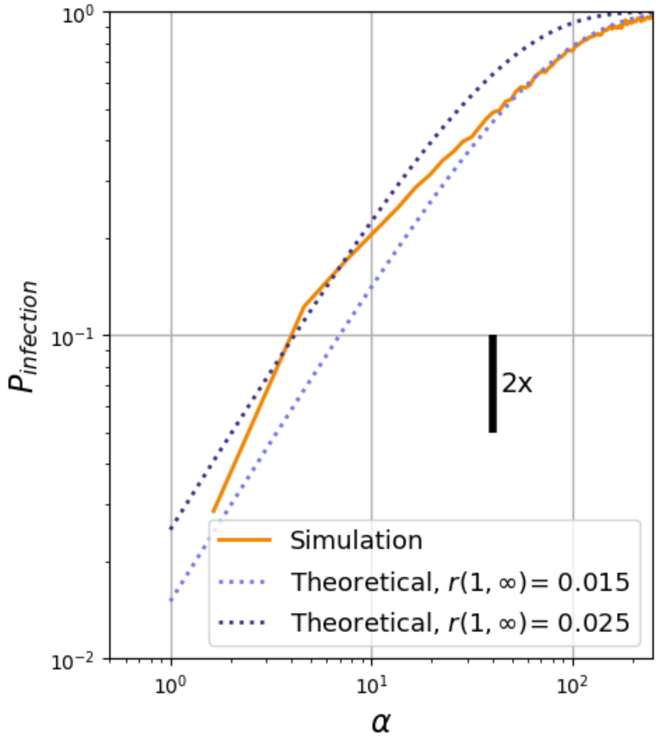


**Fig. 1.** Comparison of the probability of infection during an unmitigated epidemic in the model vs. the theoretical form predicted for random heterogeneous mixing with no network structure. The $y$-distance corresponding to a doubling is shown as a vertical bar. The light-blue curve aligns well at high activity rates, while people with $< 10$ partners per year are half as likely to get infected as in the model. The dark-blue curve fits well at low $\alpha$, but leaves those with medium $\alpha$ (30-100 partners/year) at too high risk. This shows that taking network structure into account skews the degree distribution among mpox infected significantly towards low degrees, and this allows us to obtain a good fit to the data shown in Fig. 3 A.

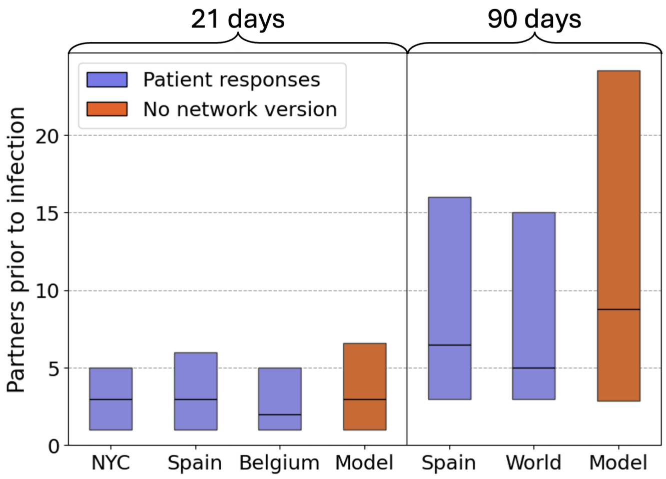


**Fig. 2.** In a variant of the model that does not take repeated encounters into account, the model cannot reproduce degree of cases on both the 21-day and 90-day timescale. Quartiles are means from 30 runs of the model.

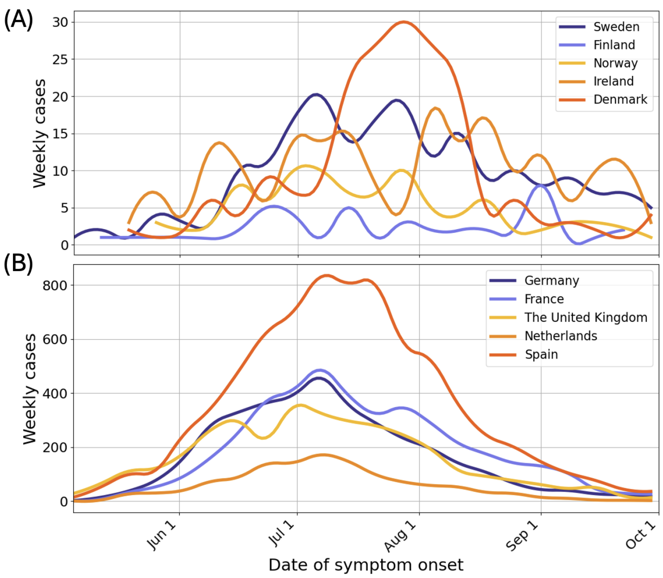


**Fig. 3.** Epidemic curves for a number for **(A)** small countries ($\lessapprox 10$ million) and **(B)** large countries. By comparison, the former are highly noisy and irregular, illustrating the founder effects from few seeds in the period before responsive behavior change.


3. T Ward, R Christie, RS Paton, F Cumming, CE Overton, Transmission dynamics of monkeypox in the united kingdom: contact tracing study. *BMJ* p. e073153 (2022).
4. M Xiridou, et al., The fading of the mpox outbreak among men who have sex with men: A mathematical modelling study. *The J. Infect. Dis.* (2023).
5. CE van Ewijk, et al., Mpox outbreak in the netherlands, 2022: public health response, characteristics of the first 1, 000 cases and protection of the first-generation smallpox vaccine. *Eurosurveillance* **28** (2023).
6. M Frisch, E Moseholm, M Andersson, J Bernhard Andresen, C Graugaard, *Sex in Denmark. Key Findings from Project SEXUS 2017-2018.* (Statens Serum Institut), (2019).
7. V Andreasen, Dynamics of annual influenza a epidemics with immuno-selection. *J. Math. Biol.* **46**, 504–536 (2003).
